# Nationwide Epidemiological Analysis of Surgically Treated Upper Limb Vascular Trauma Over 16 Years in Brazil

**DOI:** 10.1101/2024.08.12.24311874

**Authors:** Marcella Moura Ceratti, Carolina Carvalho Jansen Sorbello, Marcelo Fiorelli Alexandrino da Silva, Marcelo Passos Teivelis, Nelson Wolosker

## Abstract

**Background:** Upper limb vascular trauma (ULVT) is a prevalent injury associated with significant morbidity and mortality. Despite its clinical importance, epidemiological studies on ULVT are scarce, particularly in developing countries where the incidence may be heightened by factors such as traffic accidents and violence.

**Objective:** To analyze the epidemiology of ULVT across Brazil, a developing country, evaluating incidence rates, demographic characteristics, mortality rates, days of hospitalization, and related healthcare costs.

**Methods:** A cross-sectional, retrospective analysis using data from the Brazilian public health system (SUS) over a sixteen-year period (2008-2023). The automated data extraction utilized Python-based tools to gather information on vascular trauma procedures identified by ICD-10 codes. Statistical analyses were performed to assess variations in incidence, mortality, and treatment costs across Brazilian regions.

**Results:** A total of 25,573 cases of ULVT were recorded, representing approximately 0.02% of the studied population. The majority of cases occurred in males (79.8%) with a mean age of 34.71 years, with a peak incidence in the 20-24 age group. The region in Brazil with the highest incidence of ULVT was the North (16.6 cases per 100,000 inhabitants) and the region with the lowest was the Southeast (10.7 cases per 100,000 inhabitants). The average hospital stay was 4.39 days and 92,8% of patients did not need to be admitted to an intensive care unit (ICU). Of the patients admitted to the ICU, the average length of stay was 4.52 days. Overall lethality (deaths per cases of ULVT) was 2.37%, with higher lethality observed in bilateral ULVT cases (3.81%).

**Conclusions:** ULVT is more prevalent in Brazil than in developed countries, even when adjusted for population size. However, mortality rates and hospitalization durations do not appear to differ significantly from those in developed countries.

## INTRODUCTION

Vascular trauma accounts for approximately 2% of traumas in developed countries (1), leading to increased patient morbidity and mortality. It can result in severe bleeding, ischemic limb loss and even death (2), making it important for the medical community to be aware of.

Upper limb vascular trauma (ULVT) represent a significant portion of vascular injuries, amounting to 26% of vascular traumas in the United States (1), 23% in Scotland (3), 64% in Australia (4), and 21% in New Zealand (5). Common causes of ULVT include accidents involving sharp objects, contusions, machinery accidents, and fractures (6).

Despite the significance of ULVT, there are few epidemiological studies, based on large population data, in the literature that analyze the impact of these injuries on health systems. Only six major studies were identified in four developed nations (United States, Australia, Scotland, and New Zealand). The mortality rate for patients with vascular trauma in the United States was approximately 20% (1,6) and in New Zealand, 3.6% (4). Among patients with ULVT, lethality was 9% in Scotland (3) and 2.9% in the United States (1). Vascular injuries were more common in males, approximately 80% (2), and the average age was around 35 years (3).

Despite the relevance of these studies, all data were obtained in developed countries and cannot be extrapolated to other regions. Therefore, literature is scarce regarding the epidemiology of vascular limb traumas in large populations, particularly in developing countries where there is a higher incidence of traffic accidents and violence (7). Furthermore, no study with a large population has evaluated the cost of ULVT to the healthcare system.

Brazil had a population of 203.080.756 individuals in 2022 (8). Anyone in Brazilian territory, with any health complaint, can be attended by the Brazilian public health system (“Sistema Único de Saúde” - SUS); however, not the entire Brazilian population relies exclusively on this system (71% of the population depends exclusively on SUS) (9). SUS is considered the largest health system in the world, as no other system serves more than 200 million people (10). Data on public health in Brazil, including information about surgical procedures performed by SUS, is centralized on a portal managed by the Department of Information and Informatics of the SUS (DATASUS) (11). This system, overseen by the Secretariat of Digital Health and Information, encompasses various health-related data which includes health indicators, healthcare services, epidemiological and morbidity statistics, information on the healthcare network, vital statistics, demographic and socioeconomic data, financial details and public health expenditures. The data is anonymized and can be accessed via the online DATASUS platform, the foundation for data collection in this study.

This study aimed to evaluate the epidemiology of ULVT across Brazil, the largest country in South America and a developing nation. The report presents data for each region of Brazil on the incidence of ULVT, as well as demographic information such as gender, age group, mortality and hospitalization data including length of stay in hospitals, intensive care units and healthcare system cost.

## METHODOLOGY

### Study Period, Data Source, and population

This cross-sectional, retrospective study collected data from DATASUS. The retrospective analysis focused on procedures performed for vascular traumas of the upper limbs from 2008 to 2023. The average Brazilian population between 2008 and 2022 was approximately 196,918,278 people (8). Since 71% of the population uses exclusively the Brazilian public health system (9), the sample population for this study is at least 140,000,000 patients. However, the sample size may be much larger, since many traumas are initially treated in the SUS, so the study population is likely to be larger (between 140 and 200 million people).

### Ethical Approval

Approval for this study was obtained from the Research Ethics Committee of the institution (CAAAE 35826320.2.0000.0071). DATASUS serves as a public repository of anonymous data, eliminating the need for informed consent forms.

### Automated Data Extraction Process

The information-gathering process was automated using a Python-based (v. 2.7.13; Beaverton, OR, USA) extraction protocol devised by the institution’s IT department. Operating within the Windows 10 system, data segregation and refinement tasks were executed utilizing Selenium WebDriver (v. 3.1.8; Selenium HQ) and Pandas (v. 2.7.13; Lambda Foundry, Inc. and PyData Development Team, NY, USA) tools on the DATASUS platform.

### Procedure Selection on the Platform

First, we selected data on traumas based on ICD-10 codes (S00-S99 and T00-T98). Subsequently, we identified those who underwent the procedures “upper limb revascularization,” “surgical treatment of bilateral upper limb traumatic vascular injuries” and “ surgical treatment of unilateral upper limb traumatic vascular injuries” with DATASUS codes 0406020426, 0406020523, and 0406020531 respectively. Thus, only data regarding vascular injuries of the upper limbs resulting from traumas were extracted. In this study, ULVT treated non-surgically were not considered.

The extracted data provided information on the number of annual medical procedures, categorized by region, patient demographics (age and gender), length of hospital stay, duration in the intensive care unit (ICU), mortality rates, and financial reimbursements. Monetary values in Brazilian Reais (R$) were converted into U.S. dollars (U$) using the median exchange rate from 2008 to 2023, which was U$1 = R$ 3,2018 (12).

### Data Compilation and Statistical Analysis

Collation and structuring of data transpired in .cvs format, with Microsoft Office Excel 2019 (Redmond, WA, USA) employed for tabular arrangements. The country’s population by age group was extracted from the Brazilian Institute of Geography and Statistics (IBGE) (8). For the statistical analysis, we used the population dependent exclusively on the Brazilian public health system, which corresponds to 71% of the total population (approximately 140 million people), according to the national health agency (ANS), actualized yearly (9).

Statistical analysis was performed utilizing SPSS version 20.0 for Windows (IBM Corp, Armonk, NY). Linear regression was used to examine the progression of upper limb vascular trauma across different age groups. The chi-square and likelihood ratio tests were employed to assess the variations in upper limb vascular trauma rates among different macro-regions of the country and mortality rates within regions. A significance level of P ≤ 0.05 was deemed statistically significant for all analyses.

## RESULTS

From 2008 to 2023, there were 25,573 ULVTs, corresponding to approximately 0.02% of the study population. Out of these cases, 24,124 (94.3%) were unilateral vascular traumas, and 1,392 (5.7%) were bilateral. **Figure 1** presents the number of ULVTs per year in Brazil. There were no significant variations in the incidence of ULVTs over time. Unilateral ULVT slightly increased, while bilateral ULVT cases slightly decreased from 2008 to 2023.

**Figure 1:**
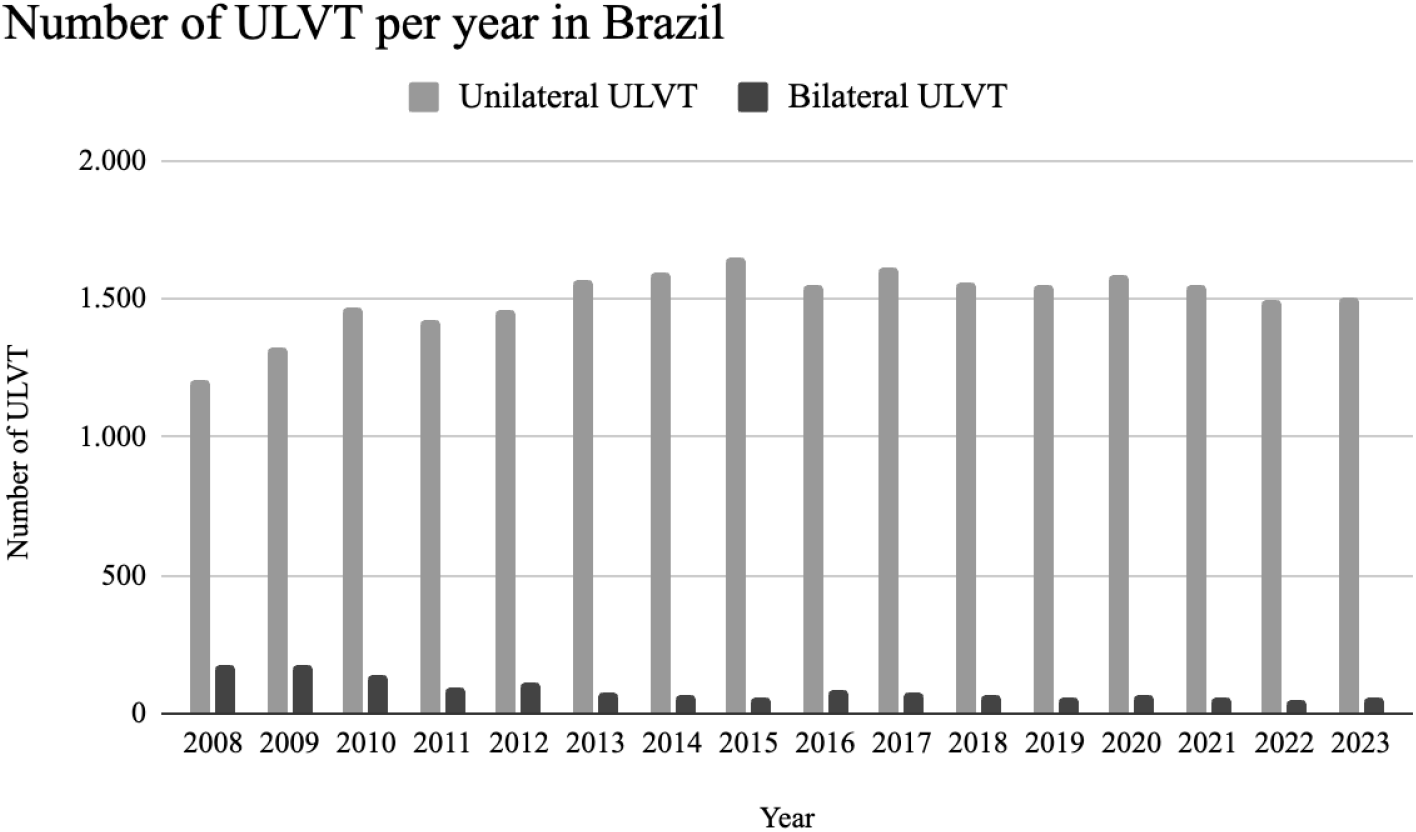
Number of ULVT per year in Brazil

**Figure 2** presents the distribution of ULVT across the five Brazilian regions. The majority occurred in the Southeast (35.4%), followed by the Northeast (30.6%). However, when compared to the populations of each region in Brazil, the Southeast had the lowest number of ULVT cases per inhabitant (despite the higher absolute number of cases), with approximately 10.7 cases per 100,000 inhabitants. The region with the most cases per inhabitant was the North (16.6 cases per 100,000 inhabitants), followed by the Northeast (14.3 per 100,000 inhabitants), the Midwest (13.0 cases per 100,000 inhabitants) and the South (12.4 cases per 100,000 inhabitants).

**Figure 2:**
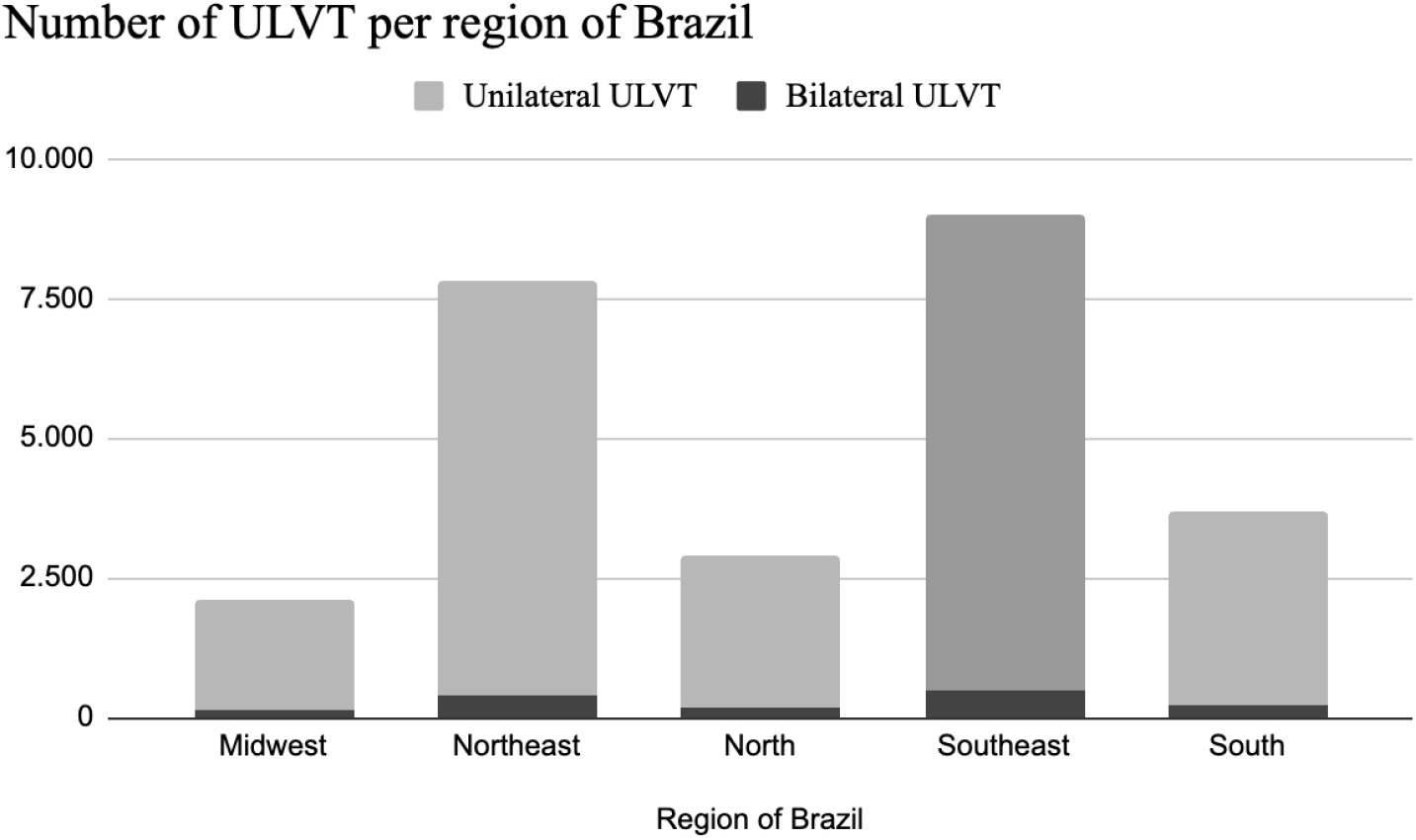
Number of ULVT per region of Brazil

**Figure 3** shows the distribution of ULVT by biological sex. Males accounted for 79.8% of ULVT cases. The bilateral/unilateral ULVT proportion in males and females was similar, at approximately 6.1% and 5.7%, respectively.

**Figure 3:**
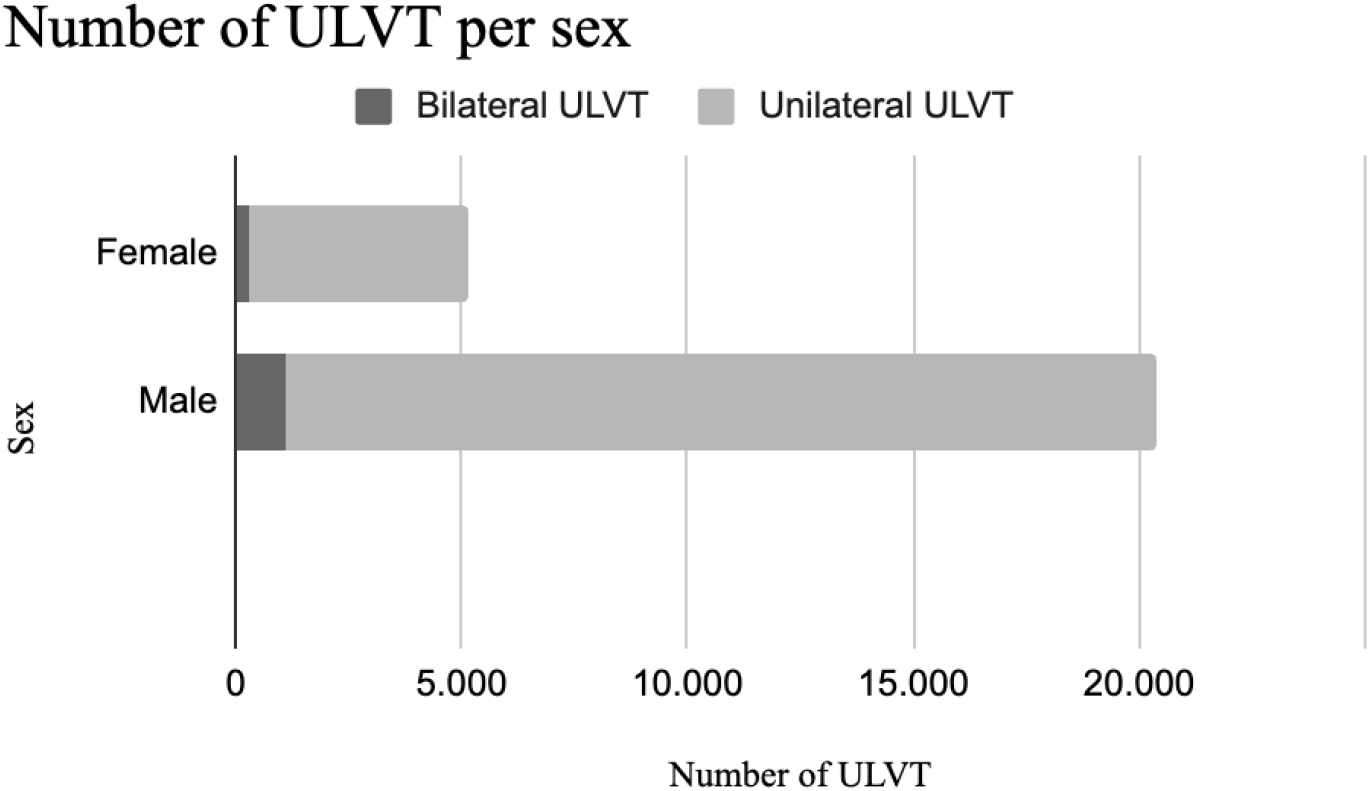
Number of ULVT per sex.

The ULVT per age group is presented in **Figure 4**. There is a peak in the incidence of ULVT at 20 to 24 years of age, with a gradual reduction as age increases. The mean age was 34,71 years.

**Figure 4:**
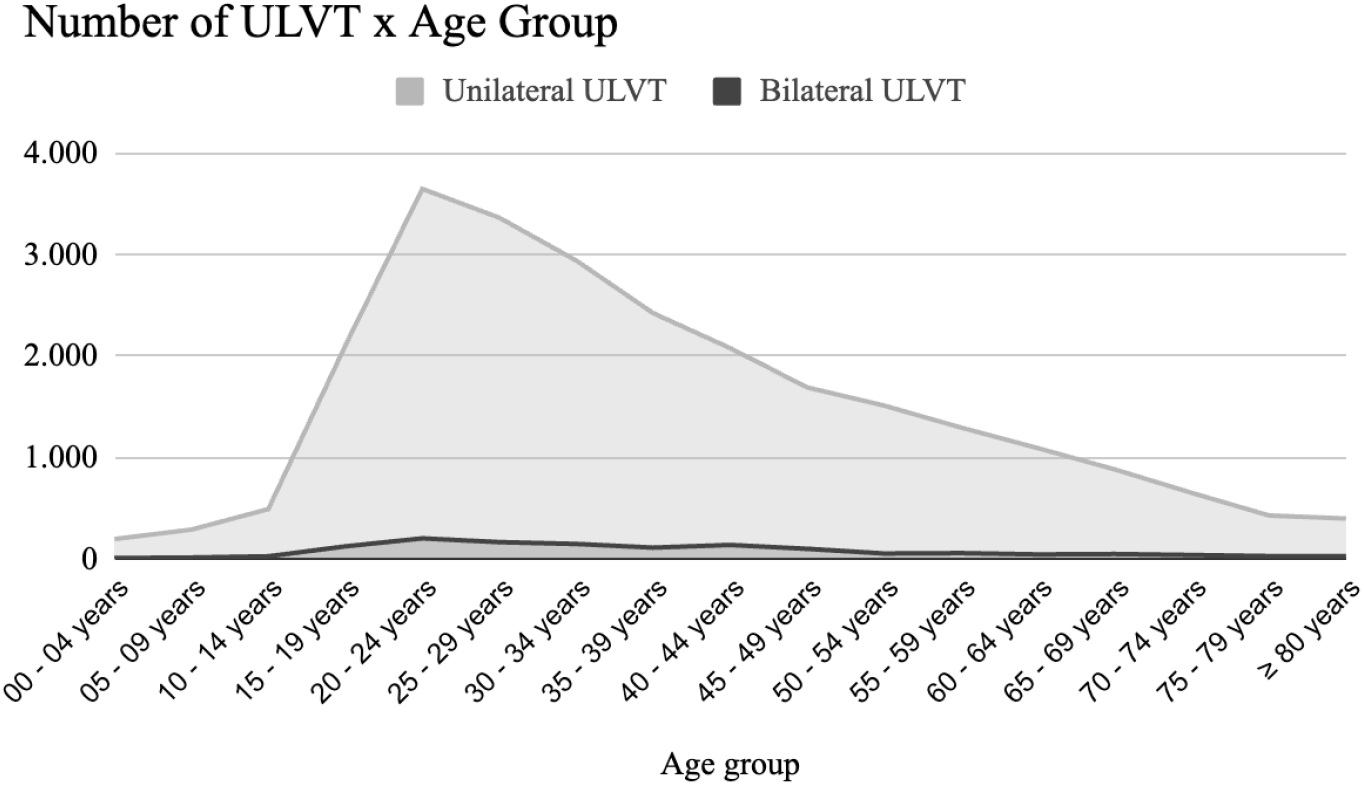
Number of ULVT per age group.

**Figure 5** shows the number of days of hospitalization. The majority of patients with ULVT were hospitalized for 2 days, with a gradual decrease in the number of days over time. The average number of days in hospital for patients with ULVT was 4,39 days.

**Figure 5:**
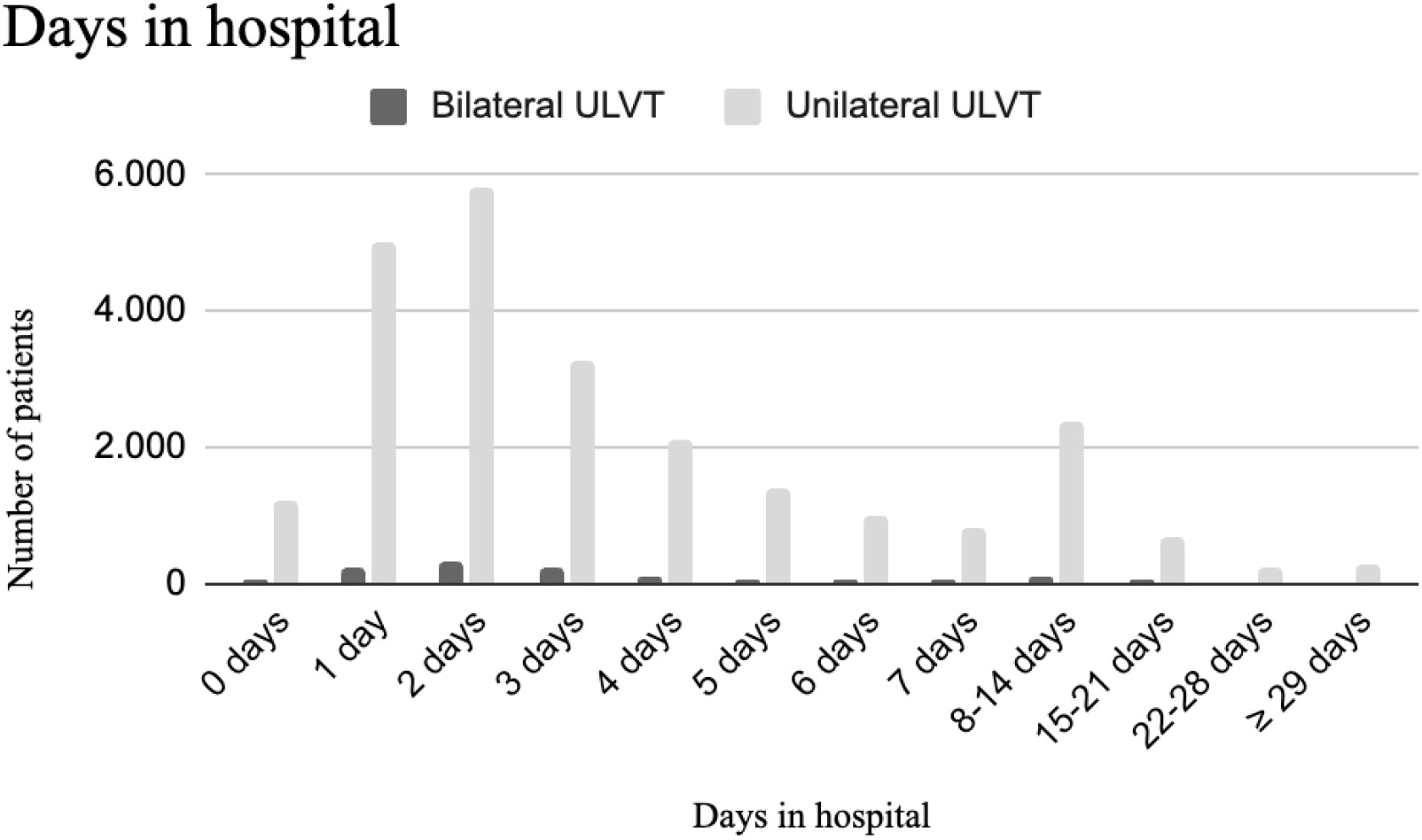
Days of hospitalization of ULVT

**Figure 6** shows the length of ICU hospitalization using a logarithmic scale. The majority of patients (92,8%) did not require ICU admission. In the group of patients that needed ICU treatment, the average number of days in the ICU was 4,52 days.

**Figure 6:**
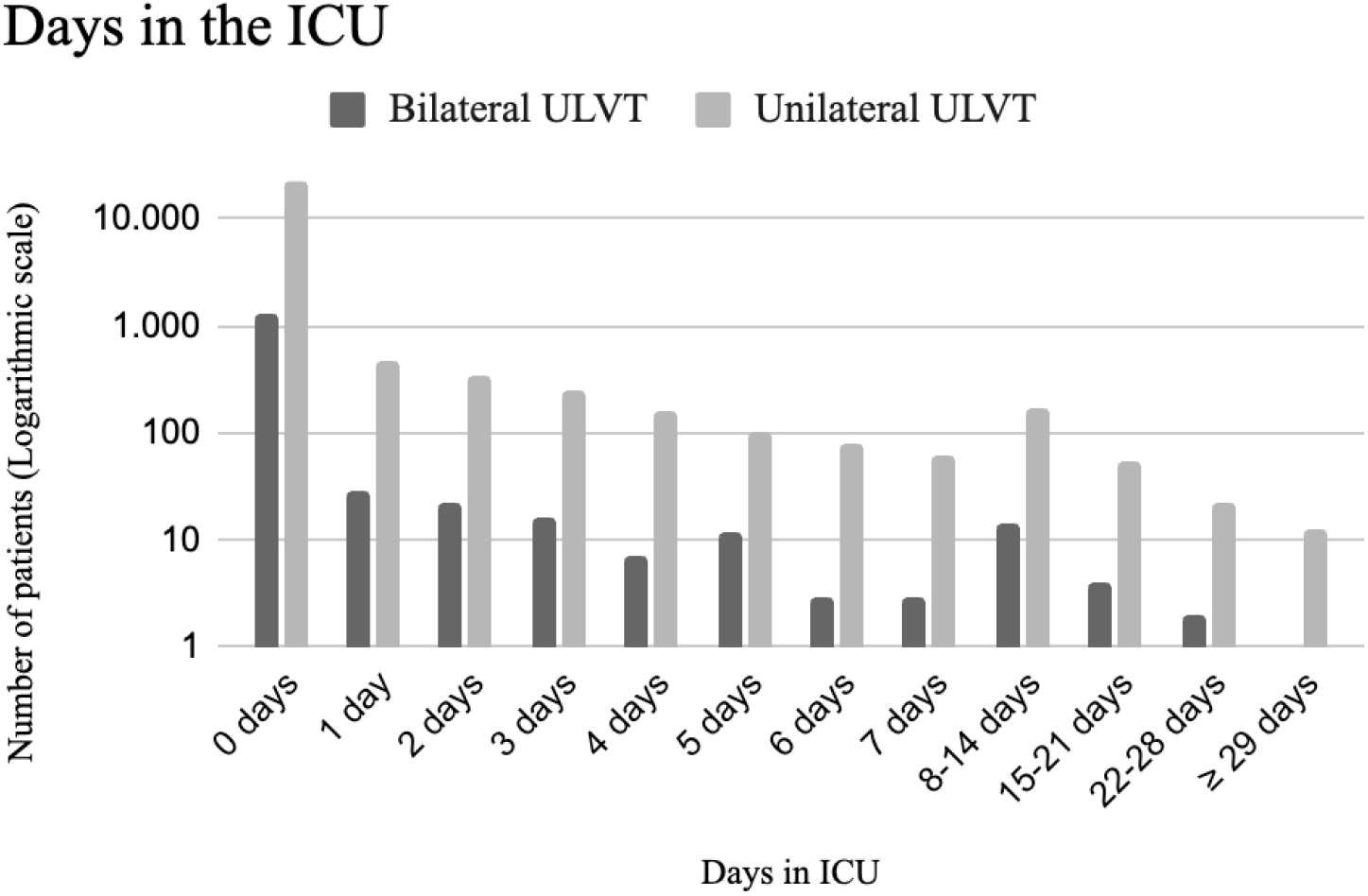
Days in the ICU for patients with ULVT.

The total number of deaths among patients with ULVT was 605, representing a lethality (deaths / cases) of 2.37%. The lethality in bilateral ULVT was higher than in unilateral cases (3.81% and 2.28%, respectively). **Figure 7** presents the lethality of unilateral and bilateral ULVT in the five regions of Brazil. It is observed that the Northeast had the highest mortality rate at 2.64%, followed by the Center-West at 2.46%. The Southeast region had the lowest mortality rate for ULVT cases, with no significant difference between bilateral and unilateral traumas (bilateral 2.77% and unilateral 2.17%).

**Figure 7:**
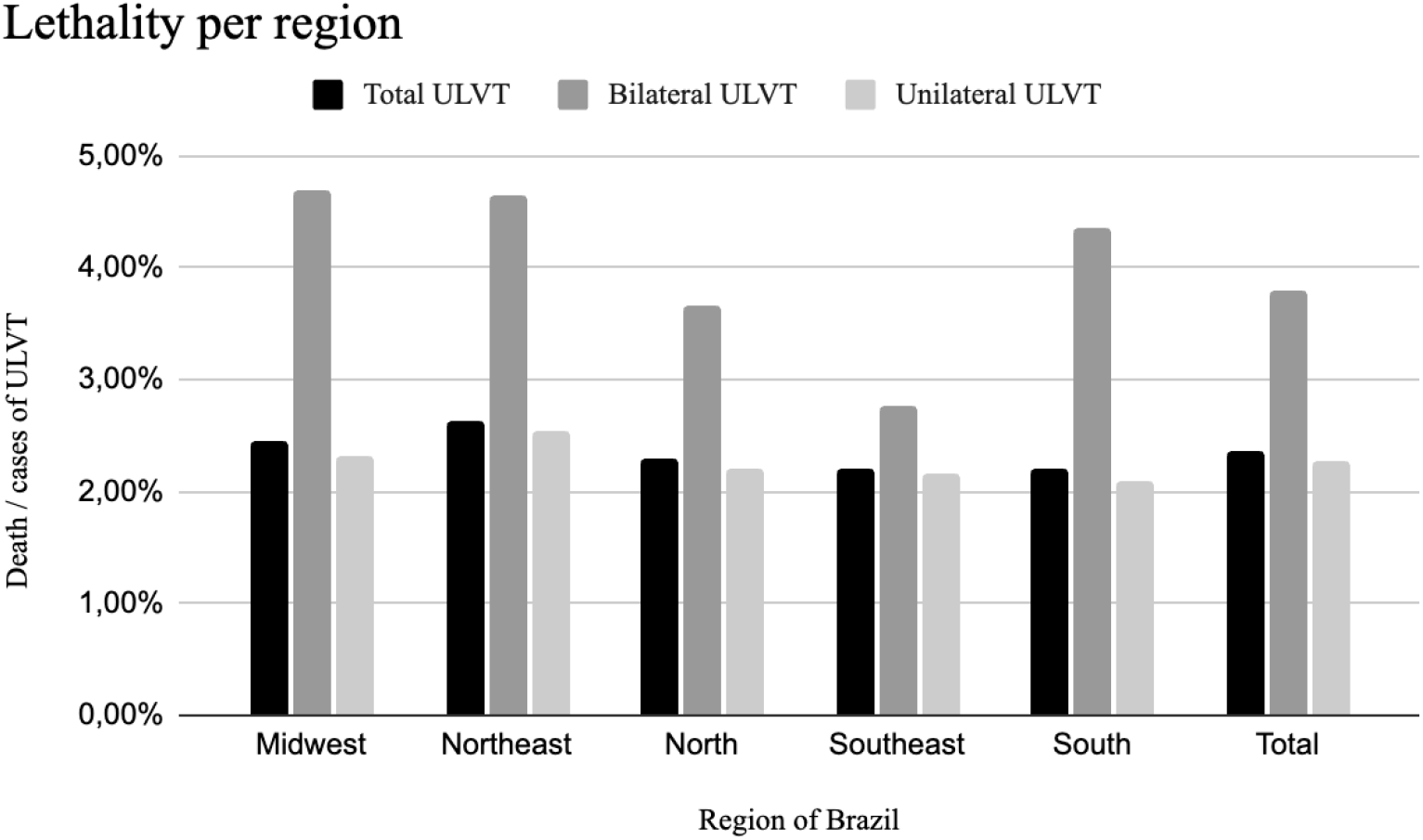
Lethality of ULVT per region of Brazil

**Figure 8** shows the distribution of lethality over the years of the study. The lethality varied considerably over the years. The year with the highest number of deaths per case of ULVT was 2020, followed by 2008 and 2010.

**Figure 8:**
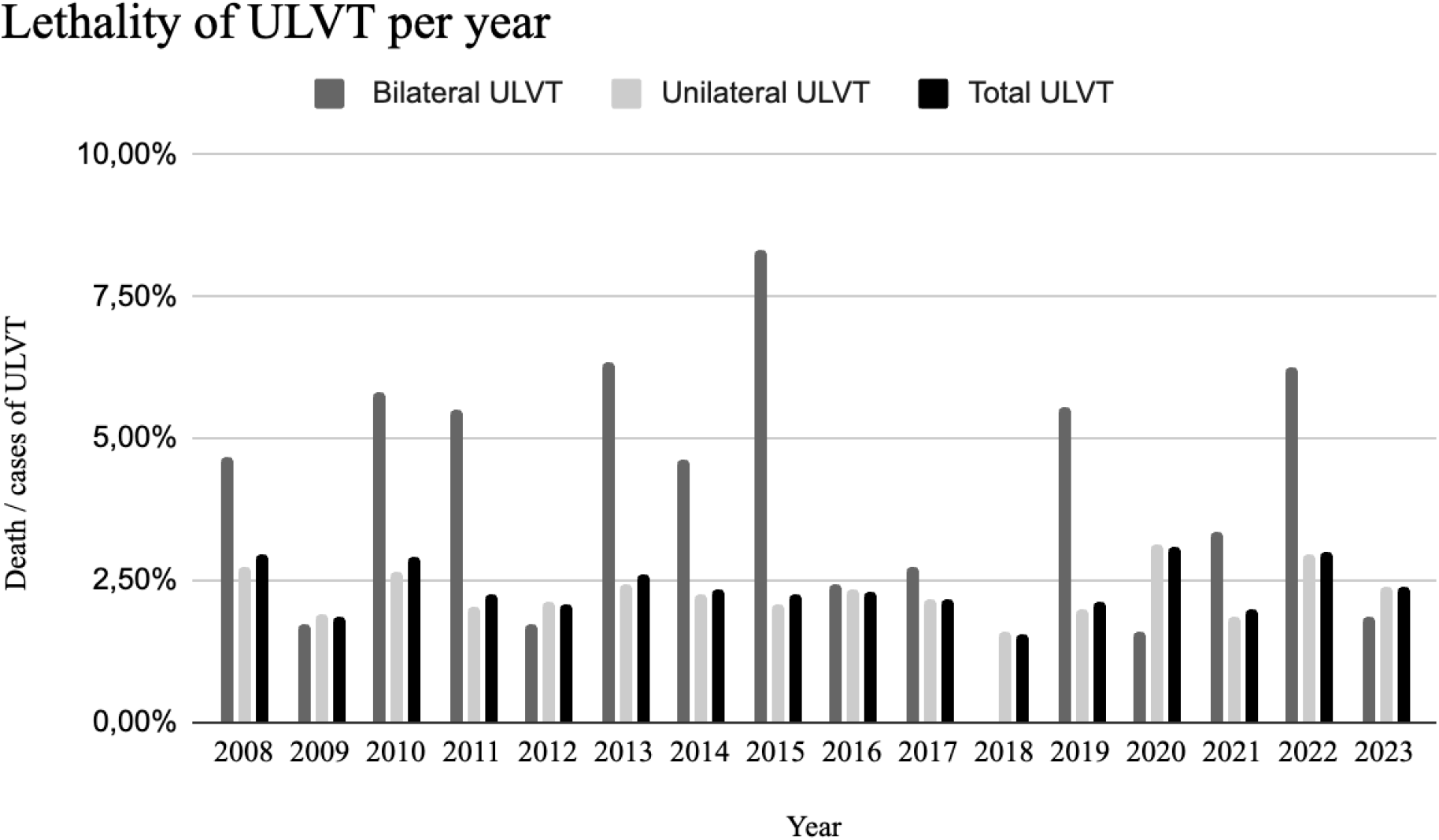
Lethality of ULVT over the years of study.

The cost for the public health system to treat all UVLT from 2008 to 2023 was $8,962,996. **Table 1** shows the total expenses by region in Brazil. The Southeast region spent the most, followed by the Northeast, which is consistent with the number of ULVT in these locations.

**Table 1:** Cost of treatment for ULVT in each region of Brazil (in dollars).

|  | Midwest | Northeast | North | Southeast | South | Total Cost | Cost / patient |
| --- | --- | --- | --- | --- | --- | --- | --- |
| **ULVT** | 734.387 | 2.839.172 | 981.417 | 3.079.147 | 1.328.873 | 8.962.996 | 350 |
| **Bilateral ULVT** | 41.575 | 131.461 | 48.886 | 181.169 | 89.311 | 492.401 | 354 |
| **Unilateral ULVT** | 691.559 | 2.704.861 | 932.531 | 2.867.163 | 1.237.669 | 8.433.783 | 350 |

**Figure 9** shows the cost per patient of treating total ULVT, bilateral ULVT, and unilateral ULVT by region in Brazil. The cost per patient did not vary much between the regions, and it is not possible to state that the treatment of patients with bilateral injuries was more expensive than unilateral ones. The South region presented a higher cost of hospitalization of patients with bilateral ULVT.

**Figure 9:**
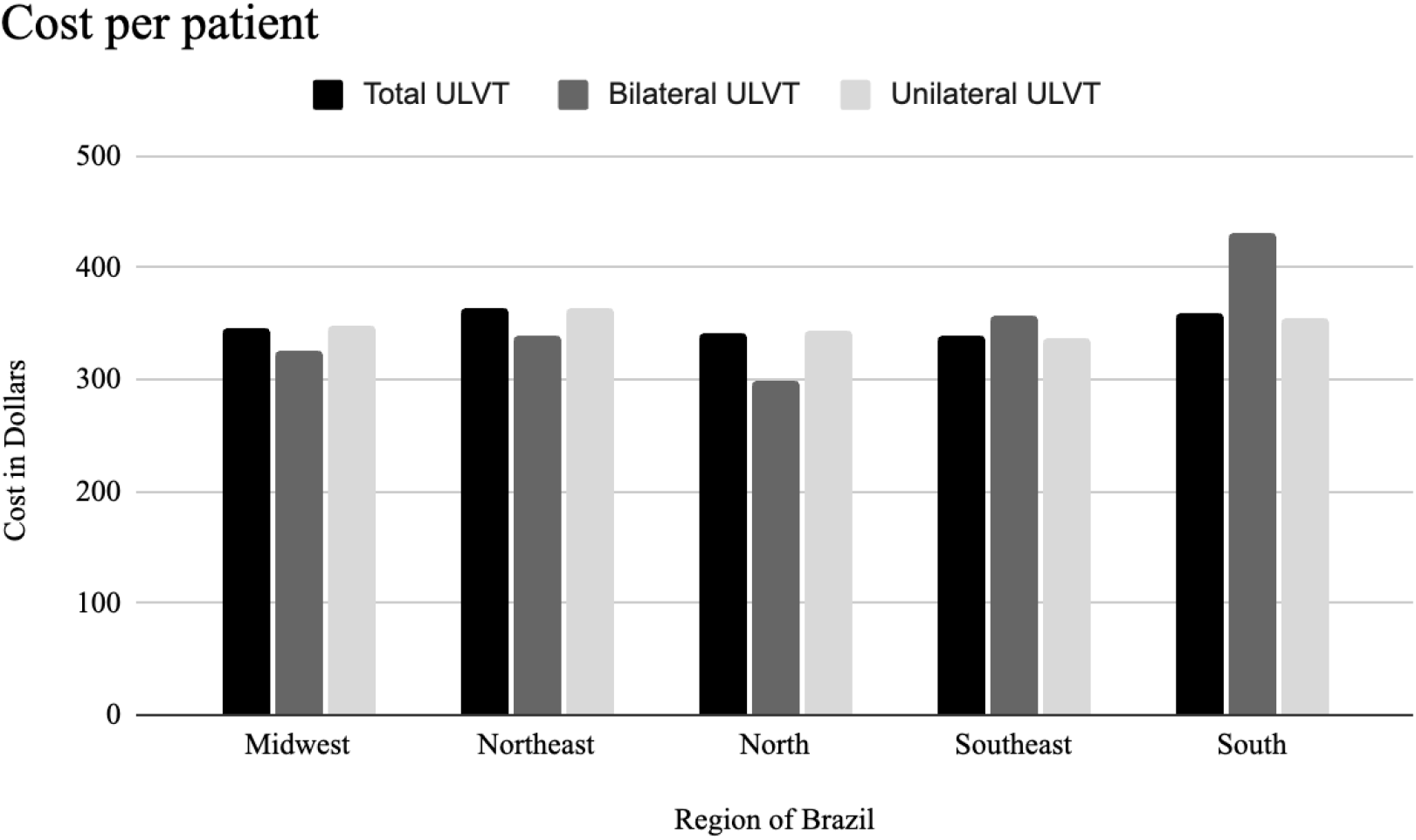
Cost per patient of ULVT (in dollars).

## DISCUSSION

Vascular trauma refers to injuries to blood vessels, which can be arterial or venous. These injuries typically result from penetrating or blunt trauma and can occur in various body regions, including the extremities, head, neck, chest, and abdomen. ULVT is one of the most common forms of vascular trauma. Treatments can vary, with traditional approaches typically involving open surgery, while endovascular methods are becoming more common (13,14). Epidemiological studies on ULVT involving large populations are scarce, particularly from underdeveloped or developing countries.

The number of ULVT in Brazil exceeds that reported in other countries studied. Despite having a population approximately 50% larger than Brazil (8,15), the United States reported fewer cases of ULVT. Over five years, Brazil documented 7,578 cases of ULVT, compared to 5,855 cases in the United States. In Scotland, a country with a population forty times smaller than Brazil, the number of ULVT cases from 2011 to 2018 was 130, while Brazil recorded 13.078 ULVT cases during the same period. Proportional to its population size, Brazil’s incidence of ULVT remains higher than Scotland’s. The higher incidence of vascular trauma in Brazil could be attributed to higher rates of traffic accidents and violence, though further studies are needed to confirm these hypotheses. Regarding the number of ULVTs over time, there has been stability in both unilateral and bilateral trauma. Not even the peak period of the Sars COV pandemic (2020-2021) has influenced the trend in cases (16).

The number of ULVT cases varied across Brazil. In absolute numbers, the Southeast reported the highest number of cases, followed by the Northeast. However, when adjusted for population size, the North had the highest incidence of ULVT cases, followed by the Northeast, Midwest, South and Southeast. These findings may be attributed to a higher prevalence of traffic accidents in the North (3.4%), followed by the Central-West (3.2%) and Northeast (2.7%), with lower rates observed in the Southeast (2.1%) and South (2.0%) (17). Additionally, physical violence seems to be more prevalent in the North and Northeast regions of Brazil, especially in areas with lower educational attainment and higher firearm availability, which may also contribute to these patterns (18).

The proportion of male patients with ULVT in Brazil was comparable to global values. In Brazil, 79.8% of ULVT cases were male, whereas in other studied countries, this number ranged from 68% to 80%. A possible explanation for these findings is that young males are at a higher risk of traffic accidents, which are related to driving behaviors and attitudes such as speeding and alcohol consumption (19). However, this remains a hypothesis, as the primary causes of ULVT in Brazil has not been definitively identified. Only one study from Latin America, which analyzed just one center in Brazil, showed that the main cause of vascular trauma, in all body regions, were firearm injuries (20).

Regarding age, the data were also consistent. Patients with ULVT in Brazil had a mean age of 34.71 years, while in the six studies, the mean age ranged from 31 to 49 years. This poses a significant issue for the labor market, as it preferentially affects an economically active age group, thereby reducing employment prospects and monthly earnings over time. Accidents and violence, along with chronic diseases, are among the leading causes of economic loss (21).

The mean length of hospital stay for patients with ULVT showed little variation among the countries studied. In the United States, two studies reported average stays of 4.6 and 5.7 days (1,6), while in Australia, one study reported an average stay of 2 days (4). Meanwhile, in Brazil, the mean length of hospital stay for ULVT patients was 4.39 days. Therefore, the duration of hospital stay was relatively consistent across the countries studied.

Concerning patients requiring intensive care, two studies from the United States showed that the duration of ICU stay for patients with ULVT averaged 4.5 and 4.6 days (1,6). These results are consistent with Brazil, where the average days in ICU was 4.52 for patients that required intensive treatment (only 7,2% of patients with ULVT).

The mortality rate for ULVT patients in Brazil was 2.37%, compared to a range of 2.2% to 2.9% in the United States (1,6), 9% in Scotland (3), and 0.9% in Australia (4). Within Brazil, the Northeast region had the highest lethality rate for ULVT patients, followed by the Midwest, North, South, and Southeast. One hypothesis for this difference is the uneven distribution of the country’s largest medical centers, which are more concentrated in the Southeast and less so in the North (22).

To date, no studies on large populations have assessed the costs of ULVT for the healthcare system. This study observed that a total of $8,962,996 was spent on the acute treatment of these injuries, representing a cost of $350 per patient. However, additional costs for post-hospitalization therapies, such as rehabilitation, follow-up consultations, and work incapacity, were not considered. These costs can be much higher when considering that these are injuries generally in economically active populations that may partially or totally lose their ability to work.

In conclusion, ULVT is more prevalent in Brazil compared to other countries, even when adjusted for population size. This suggests that underdeveloped and developing countries may experience higher rates of ULVT than developed countries. However, the mortality rate and length of hospitalization for ULVT in Brazil do not exceed those observed in developed countries. There is some regional variation within Brazil concerning the incidence and mortality of ULVT.

## LIMITATIONS

A limitation of this study is that it is not possible to determine the exact number of the sample evaluated. It is known that the sample is at least 140 million people, as this is the number of people who rely exclusively on the SUS. However, it is likely that this number is higher, since traumas are usually treated acutely by the SUS, even if the patient is later transferred to the private system. Thus, the study population could vary from approximately 140 to 200 million people.

As with any population-based study, there is always the possibility of measurement bias in some cases. This occurs because it relies on medical records by healthcare professionals, which may underestimate the number of vascular traumas, especially those involved with other traumas.

Another limitation is that we didn’t take into account medically treated ULVTs and primary amputations. The data obtained was extracted using surgical codes, which limits access to the number of clinical treatments, which may have underestimated the number of ULVTs. Despite this, limb trauma is mostly treated with surgery (23,24) which allows this amount to be extrapolated as the total number of ULVTs for analysis purposes. Based on practice, clinically treated vascular injuries are usually not recorded as vascular injuries. Most of the other studies reviewed made no distinction between what was treated clinically or surgically.

Furthermore, the costs presented in this study are the reimbursements paid to SUS based on a fixed schedule, so they may not reflect the actual cost to the hospital. Finally, one last limitation is that the data is anonymous, therefore it does not allow for follow-up on the outcomes of the patients studied.

## CONCLUSION

ULVT in Brazil is more prevalent than in developed countries and primarily affects young males. While regional variations in incidence and mortality exist, these differences are not significantly pronounced. The study underscores the need for targeted prevention strategies and calls for further research into the causes and economic impacts of ULVT in developing countries.

## Data Availability

All data produced in the present work are contained in the manuscript

https://datasus.saude.gov.br/

## Notes

### Competing Interest Statement

The authors have declared no competing interest.

### Funding Statement

This study did not receive any funding

### Author Declarations

The study used (or will use) ONLY openly available human data that were originally located at DATASUS (Department of Information and Informatics of the brazilian public health system) https://datasus.saude.gov.br/

